# Understanding the social inclusion needs of people living in mental health supported accommodation

**DOI:** 10.1101/2023.05.04.23289515

**Authors:** Sharon Eager, Helen Killaspy, C Joanna, Gillian Mezey, Megan Downey, Brynmor Lloyd-Evans

**Author notes:** Corresponding author: Prof. Brynmor Lloyd-Evans, Division of Psychiatry UCL, Maple House, 149 Tottenham Court Road, London W1T 7NF.

## Abstract

**Objectives:** To identify the social inclusion needs that were i) most commonly identified and ii) most and least commonly prioritised as support planning goals for mental health service users living in supported accommodation, using the online Social Inclusion Questionnaire User Experience (SInQUE). We qualitatively examined mental health supported accommodation staff and servicer users’ views on barriers to offering support with two less commonly prioritised areas: help finding a partner and feeling less lonely.

**Methods:** Anonymous SInQUE data were collected during a completed study in which we developed and tested the online SInQUE. Four focus groups were conducted with mental health supported accommodation staff (N=2) and service users (N=2).

**Results:** The most common social inclusion needs identified by service users (n=31) were leisure activities, finding transport options, and feeling less lonely. Of the needs identified, those that service users and staff least frequently prioritised as support planning goals were having company at mealtimes, getting one’s own furniture, feeling less lonely, help with finances, and help finding a partner. In the focus groups, staff and service users identified barriers to helping with loneliness and finding a partner which related to staff and service users themselves, supported accommodation services, and wider societal factors.

## Introduction

Social inclusion refers to an individual’s opportunity to participate as they would like in the key activities of the society where they live [1, 2]. It encompasses many important aspects of someone’s life, including health, housing, employment, leisure activities, intimate relationships, and sense of community belonging [3].

People with serious mental health problems are among the most socially excluded in society [4]. Specifically, individuals living in mental health supported accommodation, which provides community-based support to people with particularly complex mental health problems [5], are often socially excluded [6]. They frequently report feelings of loneliness, low levels of employment, and a lack of intimate relationships [7, 8]. Among those with serious mental illnesses, social inclusion is associated with better quality of life, while social exclusion is associated with poorer mental health outcomes [9, 10]. However, people with serious mental illness often report that they do not get as much support as they would like with social inclusion, and specifically social relationships [11].

The SInQUE is a measure to assess social inclusion in people with severe mental illness [12]. It has been validated across different mental health populations, has established reliability, and is acceptable to service users [9, 13]. In a recent study, the measure was adapted and developed as an online tool to inform support planning for people living in mental health supported accommodation [14]. An online SInQUE assessment is completed by staff and service users collaboratively using a computer, tablet, or other hand-held device. It produces a list of the aspects of social inclusion that the person has said they would like more support with, and up to three of these can be selected for integration into their support plan. The online SInQUE has been deemed acceptable, user-friendly, cross-culturally appropriate, and potentially useful by supported accommodation staff and service users [14].

In the current paper, we examine the areas of social inclusion that were most commonly identified and which of these were least frequently prioritised as support planning goals by staff and service users who used the SInQUE during its development [14]. We also report staff and service users’ views, elicited from focus groups, on barriers to meeting identified social inclusion needs relating to social relationships. This paper reports findings from the completed ‘SUSHI’ study (approved by the London Camden and Kings Cross Research Ethics Committee, reference: 21/LO/0657).

### Aims

We used data from SInQUE assessments completed by staff and service users of mental health supported accommodation services in one inner London borough to explore:

1. Which aspects of social inclusion were most commonly identified by service users as areas where they would like more support?
2. Which of these identified needs were most and least frequently selected by staff and service users as priorities for support planning?

We also report findings from four focus groups, two of which were conducted with mental health supported accommodation staff (N=11) and two with service users (N=7), which explored perceived barriers to offering support with finding a partner and feeling less lonely – two needs that were infrequently prioritised for support planning.

## Methods

### SInQUE data

The online SInQUE [15] was made available for use in mental health supported accommodation services across one inner-London borough, along with an implementation strategy to encourage its use. A full description of the online SInQUE and the implementation strategy employed are reported elsewhere [14].

The online SInQUE randomly generates a unique identification number for each service user using the tool, so the identity of service users who have completed an assessment is not known. Anonymous responses were collected from those using the online SInQUE from the beginning of the implementation period, starting on the 11^th^ of May 2022. Assessment responses up until the end of the study on the 30^th^ of November 2022 were collated to identify the areas of social inclusion that participating supported accommodation service users across the borough most frequently wanted help with, and those that service users and staff most frequently prioritised as support planning goals.

### Focus groups

We recruited both staff and service users who had and who had not used the online SInQUE for the four focus groups. The topic guide was informed by the collated responses from the online SInQUE. Questions related to aspects of social inclusion that were most frequently chosen as support planning priorities, and asked about potential barriers to support with areas of unmet need identified by the tool (for topic guide see Additional file 1).

We recruited staff participants through direct emails and phone calls. Two supported accommodation managers helped recruit participants for the service user focus groups by distributing the study information to individuals living in their respective services and then sending the contact details of those interested to the study team.

One staff focus group was entirely face-to-face, while one was a hybrid face-to-face and online meeting, via Microsoft Teams. Face-to-face meetings took place in the local mental health community rehabilitation team base. Both service user focus groups were face-to-face and took place onsite in each respective supported accommodation service. The study researcher discussed the information sheet with all participants at the start of each focus group, offering the opportunity for questions. Informed consent was collected with paper consent forms (audio recorded consent was collected for participants joining via video call). Focus groups were recorded using a digital voice recorder and audio recordings were transcribed by a professional transcription company. Service user participants were offered a £20 shopping voucher to thank them for their time.

For this report, our analysis focused on exploring experiences and perceived barriers to support with finding a partner and feeling less lonely – two areas of need which were rarely selected as targets for support. We used a deductive framework approach to analyse the focus groups, categorising findings based on whether they related to service users, staff, supported accommodation services, or wider society. These headings formed four primary themes, under which we inductively derived subthemes using thematic analysis. BLE developed the deductive analytic framework, and thematic coding was conducted by SE.

## Results

### SInQUE data

Of the 28 areas of life where all SInQUE respondents are asked if they would like to be more included, the participating service users said they would like to be more included for a median of 9 items. Table 1 shows the areas that service users most commonly selected as needs for greater inclusion (identified needs), and of these, the areas that staff and service users together most frequently selected as goals for support (addressed needs). Of the needs identified by at least 10 respondents, those most frequently addressed were going to a café or pub (70%) and leisure activities (58%). Five areas of need identified by at least 10 people were addressed for less than 20% of respondents: having company at mealtimes, getting one’s own furniture, feeling less lonely, help with finances, and help finding a partner.

**Table 1.** Social inclusion needs of mental health supported accommodation service users (N=31).

| Rank | SInQUE assessment areas | Identified need | Addressed need |
| --- | --- | --- | --- |
| 1 | Leisure activities | 24 | 14 (58%) |
| 2 | Get transport options | 22 | 6 (27%) |
| 3 | Feel less lonely | 19 | 3 (16%) |
| 4= | Have a holiday | 18 | 7 (39%) |
| 4= | Help with finances | 18 | 3 (17%) |
| 6= | Help to get a dentist appointment | 17 | 5 (29%) |
| 6= | Get some furniture of my own | 17 | 2 (12%) |
| 8= | Start saving money | 16 | 8 (50%) |
| 8= | Find a partner | 16 | 3 (19%) |
| 10= | See my family more | 14 | 4 (29%) |
| 10= | Reduce fear of crime | 14 | 3 (21%) |
| 12 | Help with physical health problems | 13 | 4 (31%) |
| 13= | Go to a café or pub | 10 | 7 (70%) |
| 13= | Go to a community centre or cultural organisation | 10 | 3 (30%) |
| 13= | Help getting involved in education | 10 | 2 (20%) |
| 13= | Have company at mealtimes | 10 | 0 (0%) |
| 17 | Get a computer/laptop | 8 | 2 (25%) |
| 18 | Do religious worship | 7 | 4 (57%) |
| 19= | Make friends | 5 | 2 (40%) |
| 19= | Help finding paid employment | 5 | 0 (0%) |
| 19= | Improve online access or know-how | 5 | 0 (0%) |
| 22 | Get a mobile phone | 3 | 0 (0%) |
| 23= | Get some new clothes | 2 | 2 (100%) |
| 23= | See my children more | 2 | 0 (0%) |
| 23= | Open a bank or building society account | 2 | 0 (0%) |
| 26 | Help finding voluntary work | 1 | 0 (0%) |
| 27= | Improve housing situation | 0 | 0 (0%) |
| 27= | Register to vote | 0 | 0 (0%) |

Data reported as N and N (approximate %), where N=number of participants.

### Focus groups

In total, eleven staff members participated; seven took part in the first focus group and four took part in the second. Seven service users participated overall; three joined the first focus group and four joined the second. Participant characteristics are summarised in Table 2.

**Table 2.** Demographic characteristics of staff and service user focus group participants.

| Participant characteristics | Staff (N=11) | Service users (N=7) |
| --- | --- | --- |
| **Gender** |  |  |
| Male | 8 (73%) | 4 (57%) |
| Female | 3 (27%) | 3 (43%) |
| Non-binary | - | - |
| **Age** |  |  |
| 18 – 30 | 3 (27%) | 2 (29%) |
| 31 – 50 | 3 (27%) | 2 (29%) |
| 51+ | 4 (36%) | 3 (43%) |
| Prefer not to say | 1 (9%) | - |
| **Ethnicity** |  |  |
| White/White British | 5 (45%) | 4 (57%) |
| Black/Black British | 2 (18%) | 1 (14%) |
| Asian/Asian British | 1 (9%) | - |
| Mixed/multiple ethnic groups | 1 (9%) | 2 (29%) |
| Prefer not to say | 2 (18%) | - |
| **Sexual orientation** | *N/A<sup>a</sup>* |  |
| Heterosexual/straight | - | 5 (71%) |
| Gay/lesbian | - | - |
| Bi/Bisexual | - | 1 (14%) |
| Prefer not to say | - | 1 (14%) |
| **Type of supported accommodation lived/worked in** |  |  |
| Floating outreach support | 3 (27%) | - |
| 9 to 5 supported housing | - | 3 (43%) |
| 24-hour supported housing | 7 (67%) | 4 (57%) |
| Residential care | 1 (9%) | - |
| **Length of time worked/lived in supported accommodation** |  |  |
| Less than 2 years | 2 (18%) | 4 (57%) |
| 2 – 5 years | 5 (45%) | 3 (43%) |
| 6 – 10 years | 1 (9%) | - |
| 10+ years | 3 (27%) | - |
| **Whether they completed a SinQUE assessment** |  |  |
| Yes | 4 (36%) | 3 (43%) |
| No | 7 (67%) | 4 (57%) |

Reported in the format N (approximate %), where N=number of participants.
<sup>a</sup>Staff were not asked about their sexual orientation.

Focus group respondents concurred that social relationships, specifically help with loneliness and finding a partner, were important needs. They identified a range of barriers to providing support to meet these needs, relating to staff and service users themselves, and wider service and societal factors. These are summarised in Table 3, with an illustrative quotation for each sub-theme generated through our analysis.

**Table 3.** Perceived barriers to supporting service users with developing relationships and loneliness.

| Domain | Perceived barrier to support | Illustrative quotation |
| --- | --- | --- |
| Service user-related | Negative history with personal relationships | “With finding a partner, it can be a bit more like difficult depending on how the person feels, because they might have gone through something that might set them back towards asking staff about that question.” - *Service user 2, focus group 1.* |
|  | Age | “When I first moved to London in my 20’s, I joined singles clubs and all that type of thing. It wasn’t just sort of to find a guy but also friends as well. [...] But when you’re in your 20’s it’s very different from when you’re in your 60’s.” - *Service user 4, focus group 2* |
|  | Mental health symptoms and medication use | “He wants to get out there and meet people, but he’s got quite severe mental health, quite acute mental health conditions, quite poor personal hygiene and spends a lot of time talking to himself.” - *Staff member 5, focus group 3.* |
|  | Lack of confidence navigating conversations | “Up until the beginning of 2017 I worked full-time. So I mean that was a big part of my conversation, your job and what you do and being in the rat race you know. But now I don’t have a job, I’m thinking ‘well, what do I talk about?’” - *Service user 4, focus group 2* |
| Staff-related | Concern about giving right information/advice to service user | “If anything goes wrong, then you might be held responsible for that. We don’t really want to, we can’t be there to give bad advice basically. And that’s what could happen. Because you don’t know the ins and outs of your client or the people that they meet. So there’s too much of a risk for us to be involved I think.” - *Staff member 1, focus group 3.* |
|  | Protecting service user vulnerability | “Some of the customers I’ve got do have a diagnosis and are very vulnerable. [...] If they are very vulnerable, you don’t want to be - what if I could get a partner who is quite abusive?” - *Staff member 3, focus group 3.* |
|  | Not considered part of staff role | “It’s not really in any of our job descriptions when you sign up to this, like I don’t know about you guys, but I give very questionable relationship advice to my friends, like I wouldn’t do it in a mental health service. It’s just not my job.” - *Staff member 8, focus group 4.* |
|  | Boundaries between staff and service user, not wanting to be overly personal | "I'm just thinking about like some clients, I don't think they would be happy if I asked them about their love life, like partners and stuff like that. I don't think they would like to talk about that with me." - *Staff member 9, focus group 4.* |
| Service-related | Services being understaffed and underfunded | "[The service is] understaffed, you shouldn't be understaffed, you shouldn't be underpaid and things like that, so then you can do the best you can do. Because if the staff's not happy, how are they supposed to do the best job they can do?" - *Service user 1, focus group 1.* |
|  | Restrictions on bringing visitors and visiting others | "You won't get [a partner] to come visit you though, can you? You're not allowed to get a male visitor here. [...] It's just like being a teenager again." - *Service user 5, focus group 2.* |
|  | Lack of organised, structured social activities to facilitate social contact | "If we had a rota per week, like Monday we do rockery, Tuesday we could do board games, Wednesday cook. [...] Like have a rota so you've got a plan." - *Service user 3, focus group 1.* |
|  | Strict remit of service, not prioritising aspects of social inclusion | "For me, I think sometimes the fundings are very restrictive, in terms of what you use it for. Also the fundings are more to engage them in a social setting, you know. Training, education, employment. I haven't seen any that are in terms of leisure activities." - *Staff member 11, focus group 4.* |
| Societal | Restrictions to staying outside a service (e.g., with a partner) linked to benefits and treatment orders | "I think to get housing benefit you have to spend so many nights a week at [accommodation], otherwise you're not eligible for housing benefit. [...] It's still a barrier." - *Staff member 2, focus group 3.* |
|  | Service users having little access to money and living in impoverished areas | "A lot of my clients, they go on about the areas they live in as well which is what stops them from being able to [engage in social activities]. [...] If you're in a known rough area or estate or whatever, it's harder for you to go out and do the activities that you want to do." - *Staff member 1, focus group 3.* |
|  | Stigma and discrimination | "In the community, they don't embrace people with mental health. For example, you can go somewhere in the park, and you know some of them with mental illness, sometimes you can tell. You can see people start like moving away or something like that." - *Staff member 3, focus group 3.* |

When discussing ways to help with personal relationships and loneliness, staff and service users acknowledged the complexity in discussing and addressing these issues. Both groups suggested confidence building as a potentially helpful approach for those seeking personal relationships. Service users also expressed a wish for increased opportunities for social activities, both within and outside their service. Some expressed a preference for such activities to take place in a different environment to where they live, though noted that they should remain local to the service to ensure accessibility.

### Findings in context

These findings are in line with previous research suggesting that mental health service users often receive less help than they would like with social relationships [11]. It also corroborates previous research from the Netherlands identifying intimate relationships as a consistent area of unmet need across supported housing [7] and international literature highlighting loneliness and social isolation as a priorities among mental supported accommodation service users [8].

The barriers for staff offering support to service users in developing relationships and loneliness can be usefully understood in line with the capability-opportunity-motivation-behaviour (COM-B) framework of behaviour change [16]. This COM-B model offers a useful lens through which to consider perceived barriers to supporting service users with developing relationships and loneliness in mental health supported accommodation. Our focus group findings suggest barriers relate to all three elements of the COM-B framework: i) capability (e.g. staff concerns about giving service users bad advice about relationships); ii) motivation (e.g. staff feeling this is not a core part of their job and doubting whether service users would appreciate being asked about personal relationships); and iii) opportunity (e.g. staffing levels and available funds limiting the help they can offer with social relationships).

### Implications

The current study highlights potentially important unmet needs for service users living in mental health supported accommodation. It also raises a possible need for staff training on navigating the personal relationship goals of the service users they support. The potential value of offering staff clarity on the remit of their role as it relates to service users’ personal relationships is also demonstrated. Future research should aim to implement the online SInQUE with a larger sample of mental health supported accommodation services from diverse areas to obtain more generalisable and representative data on needs for social inclusion in these services.

Future research should also investigate staff and service user perspectives on the types of support that are feasible and acceptable relating to social inclusion needs in mental health supported accommodation, and how this support can be integrated into services. In particular, research should examine staff and service user perspectives on how best to help with social relationships. More insight is needed to understand the help that service users want in relation to loneliness and help finding a partner, whether any such support is currently being provided, and if so, what this support is, and a more in-depth exploration of what the perceived barriers are in providing support with these areas. These topics should be explored throughout a wide range of mental health care settings, to assess whether this issue extends across the mental health care system.

### Limitations

This study has a number of limitations. The data collected from the online SInQUE was from a small sample of mental health supported accommodation services, all from within one inner-London borough. Due to the anonymity of the responses collected using the online SInQUE, we were unable to collect demographic information of the service users who completed an online SInQUE assessment. Therefore, results may not be representative of a wide range of people and may not be generalisable to other regions. This may also be true for the focus groups, participants of which were from the same inner-London borough.

Furthermore, the service user focus groups involved participants from only two supported accommodation services, and did not include service users from floating support or residential care facilities. Thus, views expressed in these discussions may not reflect views of service users from other supported accommodation services or service types.

It is unclear whether the areas of need identified by the SInQUE that were not prioritised for support planning were not chosen because they were considered less important to service users, or because they were more difficult for staff to offer support with. The focus group discussions and analysis also predominantly focused on unmet needs relating to loneliness and help finding a partner; however, these are only two of multiple areas that were infrequently chosen as support planning priorities by staff and service users.

## Data Availability

The quantitative data generated from this study is available from the corresponding author upon reasonable request. In line with our ethical approvals, the qualitative data generated from this study is not publicly available. This is to preserve participant anonymity.

## Abbreviations

UCL: University College London
SInQUE: Social Inclusion Questionnaire User Experience
COM-B: Capability-Opportunity-Motivation-Behaviour.

## Declarations

### Ethics approval and consent to participate

This research was conducted as part of the project: SUSHI Phase 2. This study was approved by the London Camden and Kings Cross NHS Research Ethics Committee on 04/11/2021 (REC reference 21/LO/0657).

### Consent for publication

Not applicable.

### Availability of data and materials

The quantitative data generated from this study is available from the corresponding author (BLE) upon reasonable request. In line with our ethical approvals, the qualitative data generated from this study is not publicly available. This is to preserve participant anonymity.

### Authors’ contributions

SE led recruitment, data collection, data analysis, and drafting the paper. BLE led study design, project management, and assisted with data analysis and drafting the paper. HK, JC, and GM co-designed the study and supervised the project. MD contributed to recruitment and data collection. All authors read, critically revised, and approved the final manuscript.

## Acknowledgements

We would like to thank the supported accommodation managers, staff, and service users who supported and participated in this study. We would also like to thank members of our expert advisory group and the UCL Service User Research Forum (SURF) for their advice and feedback throughout the project.

## Additional files

Additional file 1: focus group topic guides.

File format: Microsoft Word document.

