## Additional file 1 for "Understanding the social inclusion needs of people living in mental health supported accommodation"

**ADDITIONAL FILE 1: Focus group topic guides**

**Supporting social inclusion for people with serious mental illness living in supported housing: SUSHI Study Phase 2 – testing an online social inclusion assessment tool**

**Topic Guide for staff focus groups
Version 1, 31/07/2021**

*Thank you for coming to this focus group. We will ask you about ways to support people living in supported accommodation with social inclusion, and what role if any an online assessment tool can have with this. We have up to an hour for this group. It’s hard to hear what’s been said on the recording if people talk at once, so if a lot of people want to answer a question, the researcher may invite you to speak in turn, to make sure everyone gets heard. You only need to speak when you want to, and if you want to take a break or to leave at any point, that’s fine. I’ll start the recording now.*

*The research team have developed an online tool for assessing social inclusion, designed for use in mental health supported accommodation services. It’s called the SInQUE social inclusion assessment. Some of you in this group will have used the online tool and others won’t.*

**1a: For those of you who have used the online assessment: what was your experience of using it?**

Prompts:

- What led to decision to use the online assessment (own interest, encouragement from manager or colleagues? Probe impact of local implementation strategies)
- Acceptability to staff and service users (burden, usefulness)
- How did you decide when to use it and with whom? (probe cultural acceptability, timing)

**1b: For those of you who haven’t used the online assessment: what led to you deciding not to use it?**

Prompts:

- Expected acceptability or relevance to service users?
- Availability of technology?
- Concerns about using a new online tool?
- Work role and priorities?
- Lack of time?
- Prefer a less structured, or off-line conversation?

**2. What role, if any, do you think an online social inclusion assessment tool could have for day-to-day use in supported accommodation services?**

Prompts:

- Any perceived benefits?
- What, if any, are the most appropriate groups and contexts for using it?

**3. From the online assessments completed by supported accommodation staff, these areas were most commonly selected as priorities to work on together to help someone become more socially included:**

1. Leisure activities

2. Having a holiday

3. Go to a café/pub

**Why do you think that was?**

Prompts:

- Is that what you expected?
- Reasons why inclusion is difficult in these areas?
- What are some ways supported accommodation staff can help people in these areas?

**4. From the online assessments completed by supported accommodation staff, these were the areas where service users most commonly said they would like to be more socially included, but which weren’t selected as agreed priorities to work on together:**

1. Finding a partner

2. Feel less lonely

3. Help getting their own furniture

**Why do you think that was?**

Prompts:

- Difficult to increase social inclusion in these areas?
- Others are better placed than supported accommodation staff to help with these?
- Less important than other areas?

**5. Are there other ways you help people living in supported accommodation services to be more socially included which we haven’t talked about?**

**6. Are there any other resources or support you need to help people living in supported accommodation to be as socially included as they would like?**

Prompts:

- Organisational resources (time, training, managerial support or supervision, access to funds to support inclusion)
- Societal resources (more available community organisations, input from other agencies?)

**Supporting social inclusion for people with serious mental illness living in supported housing: SUSHI Study Phase 2 – testing an online social inclusion assessment tool**

**Topic Guide for service user focus groups
Version 1, 31/07/2021**

*Thank you for coming to this focus group. We will ask you about ways to support people living in supported accommodation with social inclusion, and what role if any an online assessment tool can have with this. We have up to an hour for this group. It’s hard to hear what’s been said on the recording if people talk at once, so if a lot of people want to answer a question, the researcher may invite you to speak in turn, to make sure everyone gets heard. You only need to speak when you want to, and if you want to take a break or to leave at any point, that’s fine. I’ll start the recording now.*

*The research team have developed an online tool for assessing social inclusion, designed for use in mental health supported accommodation services. It’s called the SInQUE social inclusion assessment. It has 46 questions and asks about how far someone is able to take part in different areas of life, and about areas where the person would like to be more included. Some of you in this group will have used this online tool and others won’t.*

**1a: For those of you who have used the online assessment: what was your experience of using it?**

Prompts:

- Was it clear what the assessment was for and why you were doing it?
- Acceptability (burden, anything off-putting, perceived usefulness, cultural appropriateness)

**1b: For those of you who haven’t used the online assessment: why is that?**

Prompts:

- Not been told about it by staff?
- Perceived relevance?
- Anticipated burden?
- Prefer a less structured, or off-line conversation?

**2. What role, if any, do you think an online social inclusion assessment tool could have for day-to-day use in supported accommodation services?**

Prompts:

- Any perceived benefits for service users?
- When, with whom, in what contexts could it be most helpful?

**3. From the online assessments which have been completed, these areas were most commonly selected by staff and service users as priorities to work on together to help someone become more socially included:**

1. Leisure activities

2. Having a holiday

3. Go to a café/pub

**Why do you think that was?**

Prompts:

- Is that what you expected?
- Reasons why inclusion is difficult in these areas?
- What are some ways supported accommodation staff can help people in these areas?

**4. From the online assessments which have been completed, these were the areas where service users most commonly said they would like to be more socially included, but which weren’t selected as agreed priorities to work on together with staff:**

1. Finding a partner

2. Feel less lonely

3. Help getting their own furniture

**Why do you think that was?**

Prompts:

- Difficult to increase social inclusion in these areas?
- Others are better placed than supported accommodation staff to help with these areas?
- Less important than other areas?

**5. Are there other good ways staff can help people living in supported accommodation services to be more socially included which we haven’t talked about?**

**6. Are there any other resources or support people living in supported accommodation need to be as socially included as they would like?**

Prompts:

- Other things supported accommodation services could offer?
- Support from other agencies, or from family and friends?
